# Thinking Outside the “Flow-Mediated” Box: An Analysis of Aortic Dilation in 100 Fetuses with Tetralogy of Fallot Compared to Matched Controls

**DOI:** 10.1101/2023.03.07.23286960

**Authors:** Minnie N Dasgupta, Michelle A Kaplinski, Charitha D Reddy, R Thomas Collins

## Abstract

**Background:** Aortic dilation in tetralogy of Fallot (TOF) is primarily attributed to increased aortic flow *in utero*. An alternative hypothesis is abnormal neural crest cell migration, with unequal septation of the truncus arteriosus resulting in a larger aorta and inherently hypoplastic pulmonary artery (PA). If so, we hypothesize the aorta to PA ratio (Ao:PA) in TOF is stable throughout gestation, and the total sum of dimensions of the great arteries is similar to controls.

**Methods:** We performed a single-center retrospective study of all fetuses with TOF (2014-2020) and gestational age-matched controls. We compared sums of diameters, circumferences, and cross-sectional areas of the aorta and PA and evaluated the Ao:PA across gestation in TOF with pulmonary stenosis (TOF-PS) and atresia (TOF-PA). We analyzed data with two-tailed t-tests and Pearson’s correlation.

**Results:** There were 100 fetuses with TOF (36% TOF-PA) with median gestational age of 31 weeks [IQR 26.5, 34.4] and median maternal age of 34 years [IQR 30, 37]. There were no differences in sums of great artery dimensions between TOF-PS and controls. In TOF-PA, sums were significantly lower than controls. The Ao:PA was stable throughout gestation.

**Conclusions:** The aorta in fetal TOF is large but grows proportionally throughout gestation, with a sum of great artery dimensions similar to controls. TOF-PA appears to be distinct from TOF-PS (with overall smaller dimensions), and is a group that warrants further investigation. In conclusion, our findings do not support the flow-mediated model of aortic dilation in TOF, and instead suggest an intrinsic developmental mechanism.

**Clinical Perspective:** 

**What’s New?:**

- The aorta in fetal Tetralogy of Fallot (TOF) is large, but grows proportionally throughout gestation with a total sum of great artery dimensions similar to controls.
- Fetuses with TOF with pulmonary atresia have smaller great artery dimensions than TOF with pulmonary stenosis; this distinct group warrants further investigation.
- Our findings suggest that aortic dilation in TOF may be secondary to an intrinsic developmental mechanism, rather than from increased flow to the aorta *in utero*.

**What are the clinical implications?:**

- The mechanisms of aortic dilation in fetal TOF have not been previously investigated.
- While aortic dilation is commonly seen in TOF, the degree of its progression over time and risk of dissection are not well understood.
- A better understanding of the etiology of aortic dilation in TOF could help to inform management decisions, particularly regarding the need for prophylactic surgical repair.

## Introduction

Aortic root dilation in tetralogy of Fallot (TOF) is a well-described phenomenon, from its use as a tool for identification *in utero* to late persistence following repair^1,2^. Compared to controls, patients with conotruncal defects have enlarged aortic valve and root diameters and increased aortic-to-pulmonary valve ratios^3,4^. Due to these characteristics, initial prenatal screening for TOF includes evaluating the relative sizes of the aorta and pulmonary artery^5,6^. There are multiple theories regarding the underlying mechanism of aortic dilation in TOF. The prevailing theory is flow-mediated, in which increased aortic flow *in utero* causes dilation. This is supported by data showing an inverse relationship between aortic size and size of the right ventricular outflow tract^2,7^. However, a lack of correlation between pulmonary artery and aortic sizes in other studies suggests the great arteries may not respond proportionately to changes in flow^8^. In fact, histologic abnormalities of the aorta (elastic fragmentation, medial necrosis and ground substance changes) have been reported in TOF^9,10^. These imply the aorta in TOF may be intrinsically predisposed to dilate, as in aortopathy syndromes^11–13^. Lastly, there are associations between 22q11.2 deletion and risk of aortic dilation in TOF, suggestive of a possible genetic predisposition^14,15^. Many have stressed the importance of understanding nuanced environmental and genetic contributors to congenital heart disease, as opposed to reliance on morphologic and hemodynamic models alone^16,17^. While there have been intentional investigations (with variable results) into the flow-mediated hypotheses of other lesions such as hypoplastic left heart syndrome, there has yet to be a formal study of such mechanisms in TOF^18–23^.

Thus, the objective of this study was to begin a preliminary investigation into a potential developmental etiology of aortic dilation in TOF. It is well established that the embryologic truncus arteriosus forms from neural crest cells and undergoes septation^24–26^. In TOF, neural crest cell development and migration may be altered^27,28^. Theoretically, abnormal septation causes fewer cells to migrate to the pulmonary artery, thus making it inherently hypoplastic and deviating the conal septum anteriorly. In this model, aortic enlargement in TOF results from a greater percentage of common truncal cells committing to the aorta than to the pulmonary artery. We chose to investigate the dimensional relationship of the great arteries across gestation. We hypothesized if the aorta-to-pulmonary artery ratio (Ao:PA) is stable throughout gestation, and total dimensions of the two great arteries are similar to gestational-age matched controls, then aortic dilation in TOF may be developmental, resulting from the uneven division of the common truncus arteriosus (Figure 1A). Conversely, if aortic dilation is primarily due to increased flow, longer exposure to increased flow throughout gestation should increase the Ao:PA over time and the total great artery dimensions would vary depending on individual flow ratios (Figure 1B).

**Figure 1.**
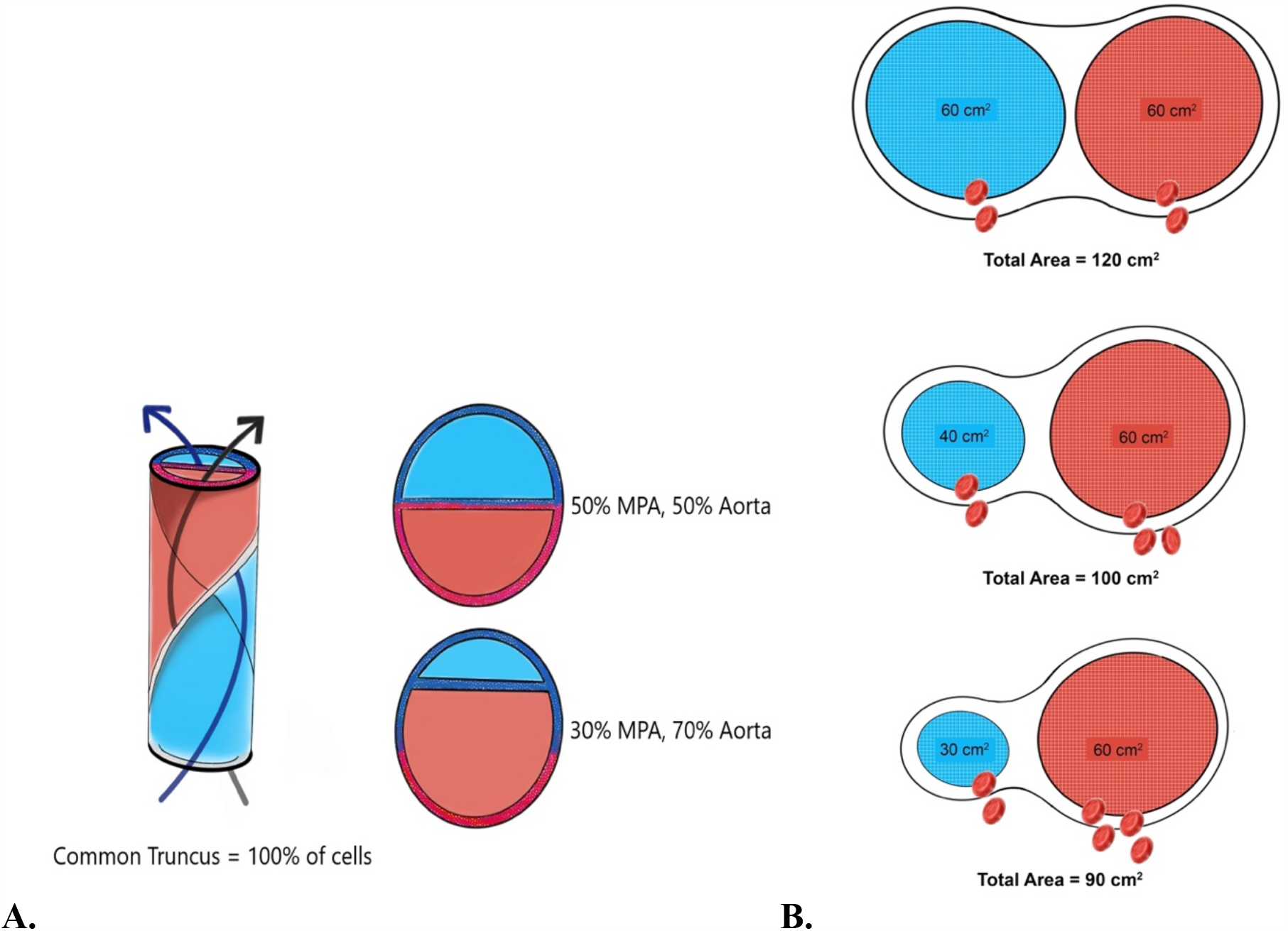
**A)** A schematic in which a common truncus arteriosus divides equally into the pulmonary artery and aorta in a normal fetus, and unequally in a fetus with Tetralogy of Fallot. The total cross-sectional area remains the same in both scenarios. **B)** A schematic depicting the expected aortic size in a flow-mediated mechanism of aortic dilation, where Qp represents blood flow through the pulmonary artery and Qs represents blood flow through the aorta. The total cross-sectional area varies with the Qp to Qs ratio.

## Methods

We performed a single-center retrospective analysis of all fetuses with TOF evaluated from December 2014 to December 2020 at the Lucile Packard Children’s Hospital Stanford. A control group was randomly selected from a large group of fetuses with normal intracardiac anatomy who were matched by equal gestational age (in weeks) and year of study. All fetal echocardiograms were stored on our institution’s secure server syngoDynamics (Siemens Medical Solutions USA; syngoDynamics Solutions, Ann Arbor, MI) with original images acquired by either an American Registry for Diagnostic Medical Sonography certified fetal sonographer or by a fetal cardiologist. If a single fetus had more than one echocardiogram available for analysis, the first and the last (chronological) studies were included as unique entries in the cohort. This was done to incorporate the possibility of changes within a single fetus across gestation. We excluded all fetuses with absent pulmonary valve syndrome, severe pulmonary insufficiency, “TOF type” double-outlet right ventricle, TOF-associated with atrioventricular canal defects or other forms of congenital heart disease, and those in whom essential cardiac structures could not be appropriately visualized. All fetuses had confirmatory postnatal imaging solidifying the fetal diagnosis, and studies were excluded when postnatal imaging was not available or if the pregnancy was terminated. The final group consisted of 100 fetuses with TOF and 100 matched controls. Of note, all control echocardiograms came from unique fetuses. The reported presence of pulmonary atresia in TOF is 10-20% in the general TOF population, but was notably high (36%) at our institution^29,30^. We therefore decided to analyze TOF in the form of three cohorts, based on severity of right ventricular outflow tract obstruction. The first cohort was fetuses with TOF with pulmonary stenosis (TOF-PS) (n = 64), the second cohort was fetuses with TOF with pulmonary atresia (TOF-PA) (n =36), and the third cohort was all fetuses with TOF, including TOF-PS or TOF-PA (n = 100).

The primary investigator (MND) made all offline measurements using the syngoDynamics workstation with 20% of randomly selected studies independently measured by the second author (MAK) for interobserver agreement. The second reader was blinded to initial analysis and subsequent interobserver variance was within 10% for all measurements. Echocardiographic measurements for this study included diameters of the aortic valve annulus, pulmonary valve annulus, ascending aorta, and main pulmonary artery. The ascending aorta was measured from both the transverse left ventricular outflow tract view and the sagittal aortic arch view, with the larger of the two measurements included for interpretation^31^. When possible, the ascending aorta was measured at approximately the level of the takeoff of the right pulmonary artery. The main pulmonary artery measurement was made just distal to the pulmonary valve when visualized. In fetuses with TOF-PA, our investigators still measured a pulmonary valve annulus size (at the valve hinge points) and branch pulmonary artery size if any valve tissue or branch pulmonary arteries, whether confluent or non-confluent, were appreciated. Patients with no appreciable pulmonary valve on fetal imaging were included in the study with a pulmonary valve annulus diameter of zero if postnatal imaging also confirmed the lack of pulmonary valve tissue. Lastly, the sex, maternal age, gestational age, biometry, and postnatal diagnosis were obtained both from the secure server and corroborated with data from the electronic medical record. Maternal race/ethnicity was also reported, in the specific categories offered for patients to select at our individual institution.

Raw data in the form of primary echocardiographic measurements were expressed in centimeters (cm). To provide multiple measures of great artery size, we calculated the sums of the diameters (cm), circumferences (cm), and cross sectional-areas (cm^2^) of the aortic and pulmonary valves, as well as the ascending aorta and main pulmonary artery. Given the normal distribution of data, we used Student two-tailed *t-*tests with equal population size and variance to compare the total great artery parameters between fetuses with TOF and normal controls. This analysis was performed for the TOF-PS group (n = 64), the TOF-PA group (n = 36), and then for the TOF-PS and TOF-PA combined group (n = 100). A p-value of < 0.05 was considered statistically significant to reject the null hypothesis that the total great artery parameters were equal. We also calculated the Ao:PA at both the valvar and proximal great artery levels. To assess these ratios over gestational age we used the Pearson correlation coefficient (Pearson’s *r*) to measure linear correlation, in which a value of 0 implies no linear dependency between two variables. Author MND had full access to all data in the study and attests to its integrity and the data analysis. The protocol for this study was approved by Stanford University’s Institutional Review Board Panel on Medical Human Subjects (protocol #50981).

## Results

One hundred fetal TOF echocardiograms and 100 normal fetal echocardiograms met inclusion criteria and made up the study cohort. The median gestational age was 31 weeks [IQR 26.5, 34.4] in both groups. There were no significant differences in baseline sex distribution, maternal age, or gestational age between groups (Table 1). Among the 100 TOF studies, there were 73 individual fetuses. Sixty-four fetal echocardiograms demonstrated TOF-PS and 36 demonstrated TOF-PA.

**Table 1:** Baseline demographics. Values for categorical variables are reported as number (percentage). *For continuous variables, values are reported as median [interquartile range].

| Variable | Tetralogy of Fallot (%) | Control (%) |
| --- | --- | --- |
| No. of Studies | 100 | 100 |
| Fetal Male Sex | 56 (56) | 52 (52) |
| Maternal Race/Ethnicity |  |  |
| White/Caucasian | 49 (49) | 32 (32) |
| Asian | 25 (25) | 23 (23) |
| Hispanic/Latino | 25 (25) | 40 (40) |
| Hawaiian Native/Pacific Islander | 1 (1) | 0 (10) |
| Black/African American | 0 (0) | 3 (3) |
| Other/Unknown | 0 (0) | 2 (2) |
| Maternal Age (years) | 34 [30, 37]* | 33 [28.8, 36]* |
| Gestational Age (weeks) | 31 [26.5, 34.4]* | 31 [26.5, 34.4]* |

Raw data were collected in the form of great artery measurements for all fetuses (Table 2). As shown in Table 3, there were no differences in the mean sums of the study measurements between TOF-PS (n = 64) and controls at the valvar or proximal great artery level. The mean sums were significantly lower in TOF-PA (n = 36) compared to controls for the diameters, circumferences, and cross-sectional areas at the valvar level, and the mean sums of the diameters and circumferences at the proximal great artery level. There was no difference in the mean sums of the cross-sectional areas at the proximal great artery level. The analysis for the full TOF cohort (n = 100) mirrored the findings of the TOF-PA cohort.

**Table 2:** Great artery measurements. Values are reported as mean ± standard deviation.

|  | **Control (n = 100)** | **All TOF (n = 100)** | **TOF-PS (n = 64)** | **TOF-PA (n=36)** |
| --- | --- | --- | --- | --- |
| **Aortic Valve Size (cm)** | 0.54 ± 0.13 | 0.70 ± 0.17 | 0.66 ± 0.17 | 0.77 ± 0.15 |
| **Pulmonary Valve Size (cm)** | 0.68 ± 0.17 | 0.31 ± 0.21 | 0.44 ± 0.11 | 0.07 ± 0.13 |
| **Ascending Aorta Size (cm)** | 0.59 ± 0.12 | 0.77 ± 0.19 | 0.71 ± 0.18 | 0.87 ± 0.16 |
| **Main PA Size (cm)** | 0.71 ± 0.18 | 0.40 ± 0.20 | 0.47 ± 0.15 | 0.23 ± 0.19 |

**Table 3:** Sums of great artery parameters. Values are reported as mean ± standard deviation. A pvalue of < 0.05 was considered statistically significant to reject the null hypothesis.

| **Tetralogy of Fallot with Pulmonary Stenosis (n=64)** |  |  |  |
| --- | --- | --- | --- |
| **Outcome** | **Control (Mean ± SD)** | **TOF -PS (Mean ± SD)** | **P-Value** |
| Aortic Valve/Pulmonary Valve |  |  |  |
| Sum of Diameters (cm) | 1.17 ± 0.28 | 1.09 ± 0.26 | 0.11 |
| Sum of Circumferences (cm) | 3.68 ± 0.87 | 3.43 ± 0.82 | 0.11 |
| Sum of Cross-Sectional Areas (cm<sup>2</sup>) | 0.57 ± 0.25 | 0.52 ± 0.23 | 0.20 |
| Ascending Aorta/Main PA |  |  |  |
| Sum of Diameters (cm) | 1.24 ± 0.29 | 1.19 ± 0.30 | 0.30 |
| Sum of Circumferences (cm) | 3.91 ± 0.90 | 3.74 ± 0.95 | 0.30 |
| Sum of Cross-Sectional Areas (cm<sup>2</sup>) | 0.64 ± 0.28 | 0.62 ± 0.29 | 0.60 |
| **Tetralogy of Fallot with Pulmonary Atresia (n=36)** |  |  |  |
| **Outcome** | **Control (Mean ± SD)** | **TOF -PA (Mean ± SD)** | **P-Value** |
| Aortic Valve/Pulmonary Valve |  |  |  |
| Sum of Diameters (cm) | 1.36 ± 0.28 | 0.84 ± 0.19 | < 0.01* |
| Sum of Circumferences (cm) | 4.27 ± 0.87 | 2.64 ± 0.61 | < 0.01* |
| Sum of Cross-Sectional Areas (cm<sup>2</sup>) | 0.76 ± 0.29 | 0.50 ± 0.18 | < 0.01* |
| Ascending Aorta/Main PA |  |  |  |
| Sum of Diameters (cm) | 1.39 ± 0.27 | 1.10 ± 0.26 | < 0.01* |
| Sum of Circumferences (cm) | 4.28 ± 0.86 | 3.45 ± 0.82 | < 0.01* |
| Sum of Cross-Sectional Areas (cm<sup>2</sup>) | 0.80 ± 0.30 | 0.68 ± 0.23 | 0.08 |
| **Tetralogy of Fallot with Pulmonary Stenosis or Pulmonary Atresia (n=100)** |  |  |  |
| **Outcome** | **Control (Mean ± SD)** | **All TOF (Mean ± SD)** | **P-Value** |
| Aortic Valve/Pulmonary Valve |  |  |  |
| Sum of Diameters (cm) | 1.24 ± 0.29 | 1.01 ± 0.27 | < 0.01* |
| Sum of Circumferences (cm) | 3.88 ± 0.91 | 3.17 ± 0.84 | < 0.01* |
| Sum of Cross-Sectional Areas (cm<sup>2</sup>) | 0.64 ± 0.28 | 0.51 ± 0.21 | < 0.01* |
| Ascending Aorta/Main PA |  |  |  |
| Sum of Diameters (cm) | 1.29 ± 0.29 | 1.16 ± 0.29 | < 0.01* |
| Sum of Circumferences (cm) | 4.05 ± 0.90 | 3.65 ± 0.92 | < 0.01* |
| Sum of Cross-Sectional Areas (cm<sup>2</sup>) | 0.70 ± 0.29 | 0.64 ± 0.28 | 0.17 |

The Ao:PA in fetuses with TOF did not increase with increasing gestational age, with a Pearson correlation coefficient *r* of 0.08 (Figure 2A). Likewise, the ascending aorta to main pulmonary artery ratio in fetuses with TOF did not increase, with a Pearson correlation coefficient *r* of -0.06 (Figure 2B). Those with no pulmonary valve tissue (pulmonary valve = 0 cm) and no main pulmonary artery segment (main pulmonary artery = 0 cm) were excluded from ratio calculations, to avoid the use of a denominator of zero.

**Figure 2.**
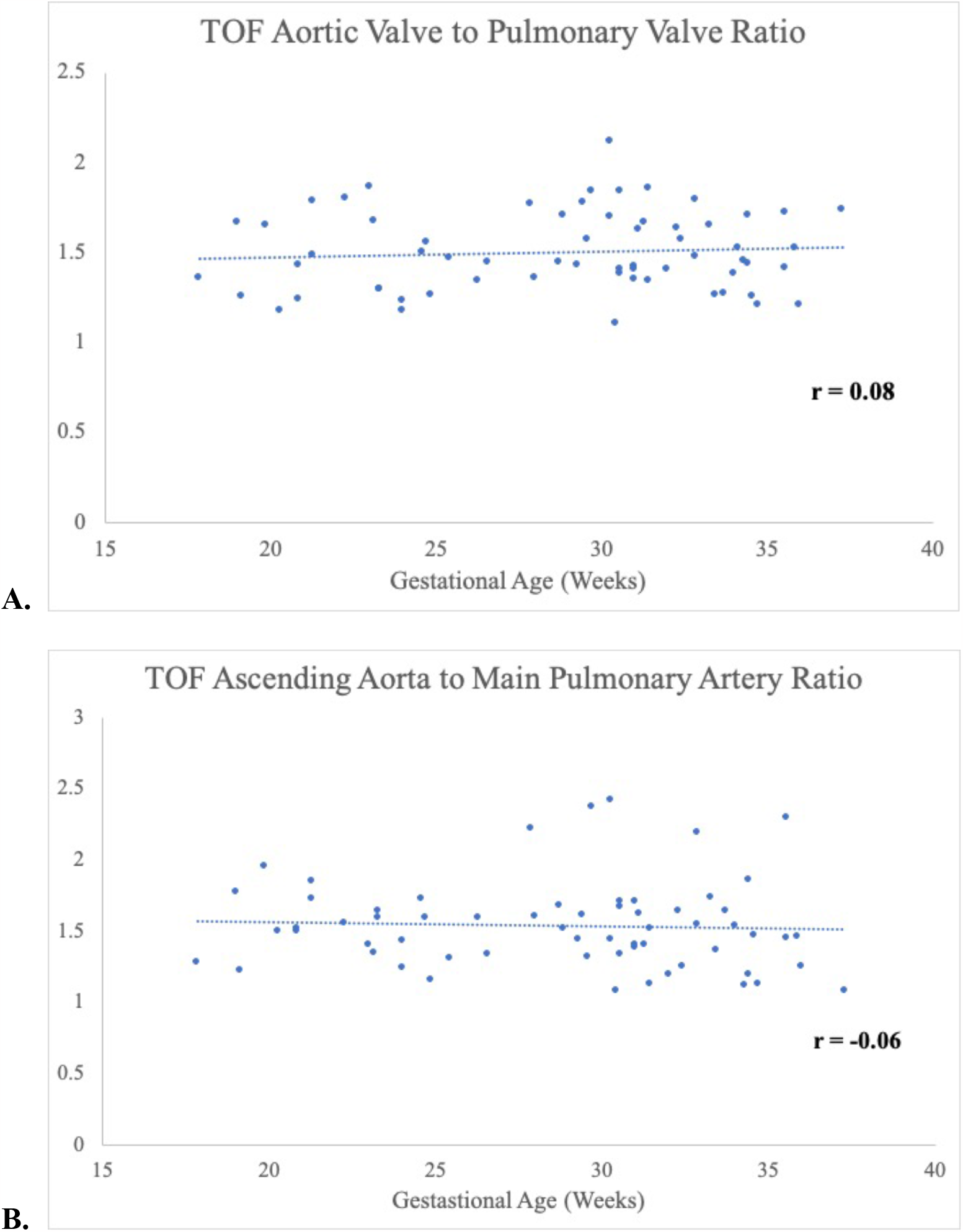
Scatter plots and associated Pearson correlation coefficients for the **A)** aortic valve to pulmonary valve ratio (*r* = 0.08) and **B)** ascending aorta to main pulmonary artery ratio (*r* = 0.06) throughout gestation in Tetralogy of Fallot.

## Discussion

This study represents one of the largest single-center cohorts of fetal TOF to date^32^. Our study sought to investigate a potential developmental etiology of aortic dilation in TOF and has three notable findings. There were no significant differences in the sums of the great artery dimensions (diameters, circumferences, and cross-sectional areas) between TOF-PS fetuses and gestational age-matched controls at the valvar or great artery level. Throughout the observed period of gestation, the Ao:PA at both measurement levels did not change. Lastly, when evaluating fetuses with TOF-PA as a unique entity, the great artery study parameters were significantly smaller than either TOF-PS or normal matched controls.

Although aortic dilation is well-described in TOF, its degree of progression and risk of aortic dissection are less well understood, with mixed data on progressive aortic changes after initial repair^1,33–36^. Overall, in large population-based studies, the risk of aortic dissection in conotruncal defects including TOF is exceedingly rare^37,38^. While catastrophic dissection in TOF is described in case reports and must be taken seriously to enable prevention, a better understanding of the etiology of aortic dilation in TOF would be helpful to explain the discrepancy in dissection rates from classic aortopathy syndromes, as well as to inform management decisions regarding the need for prophylactic surgical repair. Although this has not been previously investigated in a TOF cohort, perhaps the closest parallel theories that have been formally researched are in hypoplastic left heart syndrome, a lesion in which there are also two contrasting theories of underlying pathogenesis. Some animal studies have shown that the intentional reduction of flow across the mitral and aortic valve in fetal life can lead to the development of HLHS^18–20^. However, others have shown that there can be clinically significant left ventricular hypoplasia without associated valvar stenosis, more suggestive of a primary disturbance of cardiomyocytes mirroring a cardiomyopathy syndrome^21–23^. Similar to the argument made for understanding HLHS pathology, a stronger grasp of the etiologies of aortic dilation in TOF could better guide future investigations. To begin to understand potential contributors to aortic dilation in TOF specifically, we chose to use the sum of great artery dimensions, which is a novel method of investigating the etiology of aortic dilation in TOF. This design stems from the importance of the embryologic truncus arteriosus, which serves as the precursor to both the aorta and pulmonary artery (Figure 1A). To our knowledge, ours is the first human fetal study investigating the Ao:PA as an indicator of the septation patterns of the truncus arteriosus in TOF development. Interestingly, we found no significant differences in great artery dimension sums between TOF-PS and controls. While a flow-mediated mechanism would produce different dimension sums that vary by individual flow ratio (Figure 1B), our findings prompt the question of whether unequal truncal division in TOF may be a determinant of aortic dilation. Given this question, there remains a need for more detailed cellular analysis which could ultimately impact evidence-based guidelines on the appropriate frequency of monitoring aortic dimensions and thresholds for intervention in this cohort.

An Ao:PA greater than one is associated with TOF at any stage, but has not been previously assessed throughout gestation in such a large cohort. The general finding of enlarged aortic root dimensions compared to normal has long been an early marker for fetal diagnosis of TOF^5,39^. More specifically, an Ao:PA greater than one is frequently used for diagnosis of conotruncal defects^6,40^. Our study shows a stable Ao:PA at two defined levels (valvar and above) throughout gestation. If aortic dilation in TOF were purely flow mediated, we would expect that longer exposure to increased flow throughout gestation would subsequently increase the Ao:PA over time. The results of our study also suggest that if abnormal truncal septation plays a potential role in aortic size in TOF, an Ao:PA ratio greater than one should be apparent from quite early in gestation (theoretically, as soon as conotruncal division occurs). This second important finding again argues against the predominantly accepted flow-mediated mechanism.

As we know, TOF-PA represents the most severe form of TOF. Though the typical incidence of pulmonary atresia in TOF is 10-20%, as a large referral center for TOF-PA, our sample was skewed toward fetuses with pulmonary atresia^29,30^. Given this atypical distribution, we ultimately opted to analyze TOF in three different cohorts, as described above. Somewhat surprisingly, when including TOF-PA patients in our analysis, we found that the sums of the great artery dimensions were significantly lower than in TOF-PS fetuses and in matched controls. This was true for all parameters, with the exception of the sum of the cross-sectional areas (at the ascending aorta and main pulmonary artery level only). This unexpected finding in one parameter may be due to differences in severity of hypoplasia in the pulmonary valve as opposed to the main pulmonary artery, but warrants further consideration. In regards to the general fetal development of great arteries, it is well established that cardiac neural crest cells play an essential role in the normal development of the embryologic truncus arteriosus and its septation into the aorta and pulmonary artery. Animal models have shown that neural crest cell ablation and thus interruption of associated signaling during development leads to a broad spectrum of outflow tract abnormalities such as TOF^41–43^. The smaller dimensions seen in TOF-PA as compared to TOF-PS and normal controls is a new finding and perhaps represents a different underlying pathology altogether. Though we did not assay neural crest cell numbers in this study, one might theorize an underlying deficiency in the initial neural crest cell population that composes the common truncus in TOF-PA. If so, the common truncus may never grow to the same initial dimensions in a pulmonary atresia cohort. Since this remains theoretical, more targeted investigations are needed to better understand the molecular genetics of pulmonary atresia. However, our findings do prompt the question of whether TOF-PA may be a unique entity with a different developmental pattern from TOF with some component of antegrade flow.

There are several limitations to our study, including that it is a retrospective analysis from a single-center institution. A number of fetal studies were excluded due to poor image quality or lack of available postnatal imaging to confirm diagnosis. Standards of fetal biometry characterization also changed over the course of our study period, and data were not always available in a consistent manner from year-to-year. As a result, prior to 2016, gestational age is based on reported dates as opposed to those after 2016 which were calculated primarily on biometry; there can be significant differences between these methods of estimating age. Based on the capabilities of fetal echocardiogram, it is important to note that we were only able to assess data past approximately 18 weeks of gestation and cannot comment on changes in great artery size or ratio in earlier stages of embryologic development. In the absence of reviewing serial fetal echocardiograms for all cases in the cohort, we were also unable to specifically assess the progression of great artery sizes and ratios within each individual fetus. Other than differentiating between fetuses with and without pulmonary atresia, we did not evaluate other factors that may impact TOF physiology and variability of aortic dilation, such as streaming tendencies, degree of aortic override, or genetic conditions. Lastly, we did not evaluate post-natal outcomes or ongoing changes in great artery size in response to interventions on the right ventricular outflow tract. Several of the above limitations could be improved upon or further investigated in future versions of this study, perhaps involving serial non-invasive imaging and histology to confirm our suggested hypotheses (particularly in the postnatal period).

In conclusion, aortic dilation in TOF is well-described, but its etiology remains poorly understood with a prevailing theory of flow-mediated growth. We used a novel methodology to assess sums of great artery dimensions in utero, and found no significant differences in the sums of great artery dimensions between TOF-PS and matched controls. The Ao:PA at multiple levels also did not change throughout gestation. These findings suggest there may be a developmental component of aortic dilation in TOF, and this theory warrants further investigation.

## Data Availability

Author MND had full access to all data in the study and attests to its integrity and the data analysis.

## Acknowledgments

Sources of Funding, & Disclosures

## Statement of Financial Support

This research did not receive any specific grant from funding agencies in the public, commercial, or not-for-profit sectors.

### Declarations or Disclosures of Interest

None.

### Consent Statement

Per Stanford University’s Institutional Review Board, individual patient consent was waived for this retrospective study.

## Notes

### Competing Interest Statement

The authors have declared no competing interest.

### Clinical Trial

NA; Not a clinical trial.

### Author Declarations

The protocol for this study was approved by Stanford University's Institutional Review Board Panel on Medical Human Subjects (protocol #50981).

